# Influence of vitamin D supplementation on SARS-CoV-2 vaccine efficacy and immunogenicity

**DOI:** 10.1101/2022.07.15.22277678

**Authors:** David A Jolliffe, Giulia Vivaldi, Emma S Chambers, Weigang Cai, Wenhao Li, Sian E Faustini, Joseph M Gibbons, Corinna Pade, Alex G Richter, Áine McKnight, Adrian R Martineau

**Author notes:** Corresponding author: Blizard Institute, Barts and The London School of Medicine and Dentistry, Queen Mary University of London, 4 Newark St, London E1 2AT, UK. These authors contributed equally (David A Jolliffe, Giulia Vivaldi).

## Abstract

**SUMMARY:** *Background & Aims:* Vitamin D deficiency has been reported to associate with impaired development of antigen-specific responses following vaccination. We aimed to determine whether vitamin D supplements might boost immunogenicity and efficacy of SARS-CoV-2 vaccination.

*Methods:* We conducted three sub-studies nested within the CORONAVIT randomised controlled trial, which investigated effects of offering vitamin D supplements at a dose of 800 IU/day or 3200 IU/day vs. no offer on risk of acute respiratory infections, including COVID-19, in UK adults with circulating 25-hydroxyvitamin D concentrations <75 nmol/L. Sub-study 1 (n=2808) investigated effects of vitamin D supplementation on risk of breakthrough SARS-CoV-2 infection following two doses of SARS-CoV-2 vaccine. Sub-study 2 (n=1853) investigated effects of vitamin D supplementation on titres of combined IgG, IgA and IgM (IgGAM) anti-Spike antibodies in eluates of dried blood spots collected after SARS-CoV-2 vaccination. Sub-study 3 (n=100) investigated effects of vitamin D supplementation on neutralising antibody and cellular responses in venous blood samples collected after SARS-CoV-2 vaccination.

*Results:* 1945/2808 (69.3%) sub-study 1 participants received two doses of ChAdOx1 nCoV-19 (Oxford–AstraZeneca); the remainder received two doses of BNT162b2 (Pfizer). Vitamin D supplementation did not influence risk of breakthrough SARS-CoV-2 infection (800 IU/day vs. no offer: adjusted hazard ratio 1.28, 95% CI 0.89 to 1.84; 3200 IU/day vs. no offer: 1.17, 0.81 to 1.70). Neither did it influence IgGAM anti-Spike titres, neutralising antibody titres or IFN-γ concentrations in supernatants of S peptide-stimulated whole blood.

*Conclusions:* Among adults with sub-optimal baseline vitamin D status, vitamin D replacement at a dose of 800 or 3200 IU/day did not influence protective efficacy or immunogenicity of SARS-CoV-2 vaccination.

*Clinical Trial Registration:* ClinicalTrials.gov NCT04579640.

## 1. Introduction

Vaccination against SARS-CoV-2 represents the mainstay of COVID-19 control. However, vaccine efficacy and effectiveness wane significantly within 6 months, particularly among older adults [1]. Identification of immunomodulatory adjuvants with potential to augment SARS-CoV-2 vaccine immunogenicity is therefore a research priority [2]. Sub-optimal responses to vaccination against other pathogens in older adults are causally associated with increased systemic inflammation, termed ‘inflammaging’ [3]. Increased production of inflammatory cytokines by monocytes and macrophages is a key driver of this process [4], and pharmacological inhibition of these pathways by blocking p38 mitogen-activated protein (MAP) kinase or the mammalian target of rapamycin (mTOR) pathway, has been shown to augment antigen-specific immunity [5-7].

Vitamin D is best known for its effects on calcium homeostasis, but it is also recognised to play a key role in regulation of human immune function [8]. The active vitamin D metabolite 1,25-dihydroxyvitamin D (1,25[OH]_2_D) has been shown to inhibit production of pro-inflammatory cytokines by monocytes and macrophages by targeting MAP kinase phosphatase 1 [9], to regulate the mTOR pathway [10], and to support classical T cell receptor signalling and T cell activation by inducing phospholipase C-gamma 1 in naïve T cells [11]. Sub-optimal vitamin D status, as indicated by low circulating concentrations of 25-hydroxyvitamin D (25[OH]D, the major circulating vitamin D metabolite) is common among older adults, and this associates with increased systemic inflammation [12, 13]. An experimental study has demonstrated that vitamin D supplementation significantly increased the response to cutaneous varicella zoster virus (VZV) antigen challenge in older adults with circulating 25(OH)D concentrations less than 75 nmol/L [14]. This enhancement was associated with a reduction in early inflammatory monocyte infiltration with concomitant enhancement of T cell recruitment to the site of antigen challenge.

Taken together, these findings provide a rationale for investigating whether vitamin D replacement might enhance immunogenicity and effectiveness of SARS-CoV-2 vaccination in adults with sub-optimal vitamin D status [15, 16]. Several observational studies have investigated associations between vitamin D status and SARS-CoV-2 vaccine immunogenicity, but these have yielded conflicting results: some report higher post-vaccination titres of anti-Spike antibodies in individuals using vitamin D supplements or having higher circulating 25(OH)D concentrations [17, 18], but others have yielded null findings [19, 20]. An opportunity to investigate this question using an interventional study design arose when we conducted a phase 3 randomised controlled trial of vitamin D supplements for prevention of acute respiratory infection in UK adults (CORONAVIT) [21]. The intervention period for this study coincided with rollout of SARS-CoV-2 vaccination over Winter–Spring 2020– 21, a period when sub-optimal vitamin D status is highly prevalent in the UK [22]. We therefore nested three sub-studies within the trial to investigate effects of vitamin D replacement on SARS-CoV-2 vaccine efficacy, post-vaccination titres of anti-Spike antibodies in dried blood spot eluates, and post-vaccination neutralising antibody titres and antigen-specific cellular responses to SARS-CoV-2 in venous blood.

## 2. Materials and methods

### 2.1. Study design

We conducted three sub-studies nested within the CORONAVIT randomised controlled trial [21]. Sub-study 1 (vaccine efficacy analysis) investigated the influence of vitamin D supplementation on risk of breakthrough SARS-CoV-2 infection in immunocompetent trial participants who received two doses of a SARS-CoV-2 vaccine during follow-up. Sub-study 2 (dried blood spot analysis) investigated effects of vitamin D supplements on combined IgG, IgA and IgM (IgGAM) antibody responses to the Spike (S) protein of SARS-CoV-2 measured in dried blood spot eluates. Sub-study 3 (venous blood analysis) investigated effects of vitamin D supplements on neutralising antibody and cellular responses.

Full details relating to the design and conduct of the CORONAVIT trial and post-vaccination serology studies have been reported elsewhere [17, 21]. Briefly, 6200 UK residents aged 16 years or older and participating in the COVIDENCE UK study [23] were individually randomised to receive an offer of a postal vitamin D test, followed by higher-dose (3200 IU/day; n=1550) or lower-dose (800 IU/day; n=1550) vitamin D supplementation if their blood 25(OH)D concentration was found to be less than 75 nmol/L, or to receive no offer of vitamin D testing or supplementation (n=3100), with a 1:1:2 allocation ratio. Treatment allocation was not concealed, and randomisation was not stratified. All participants who received at least two doses of a SARS-CoV-2 vaccine were invited to provide a postal dried blood spot sample for determination of combined IgGAM antibody responses to the S protein of SARS-CoV-2, as described below. A subset of 101 trial participants also provided a venous blood sample for determination of neutralising antibody and cellular immune responses to SARS-CoV-2. The trial was sponsored by Queen Mary University of London, approved by the Queens Square Research Ethics Committee, London, UK (ref 20/HRA/5095) and registered with ClinicalTrials.gov (NCT04579640) on 8 October 2020, before enrolment of the first participant on 28 October 2020.

### 2.2. Participants

Eligibility criteria for the three sub-studies were as follows. For sub-study 1 (vaccine efficacy analysis), inclusion criteria were participation in the CORONAVIT trial and receipt of two doses of a SARS-CoV-2 vaccine, with the first dose given between 16 January 2021 (i.e. at least 1 month after the start of the trial) and 16 June 2021 (i.e. the end of the trial intervention period). Exclusion criteria for sub-study 1 were self-report of taking study vitamin D capsules less than half the time during trial follow-up (intervention arms) or self-report of any intake of supplemental vitamin D during follow-up (no-offer arm); known immunodeficiency disorder; and use of systemic immunosuppressants. For sub-study 2 (dried blood spot analysis), inclusion criteria were eligibility for sub-study 1, plus consent to participate in the post-vaccination serology sub-study and availability of an anti-S titre result from a dried blood spot sample provided at least 2 weeks after administration of the second dose of a SARS-CoV-2 vaccine and before administration of a booster dose. For sub-study 3 (venous blood analysis), inclusion criteria were eligibility for sub-study 1, plus residence within a 100-mile radius of the Blizard Institute (East London), consent to participate in the post-vaccination venous blood sub-study and availability of valid neutralising antibody or cellular response data.

### 2.3. Randomisation

CORONAVIT trial participants were individually randomised by the trial statistician to either higher-dose offer, lower-dose offer, or no offer using a computer program (Stata version 14.2; College Station, TX, USA), as previously described [21].

### 2.4. Intervention

Consenting participants randomised to either intervention arm of the trial were posted a blood spot testing kit for determination of 25(OH)D concentrations in capillary blood, as previously described [24]. Those found to have a 25(OH)D concentration below 75 nmol/L were then posted a 6-month supply of capsules containing either 800 IU or 3200 IU vitamin D_3_, according to their allocation. Participants were supplied with D-Pearls capsules of either strength, manufactured by Pharma Nord Ltd (Vejle, Denmark), unless they expressed a preference for a vegetarian or vegan supplement, in which case they were supplied with Pro D_3_ vegan capsules manufactured by Synergy Biologics Ltd (Walsall, UK). Participants with 25(OH)D concentrations of 75 nmol/L or more at initial testing were offered a second postal vitamin D test 2 months after the first test: those whose second 25(OH)D concentration was found to be less than 75 nmol/L were offered a postal supply of supplements as above. Participants receiving study supplements were instructed to take one capsule per day until their supply was exhausted. Administration of study supplements was not supervised.

### 2.5. Follow-up assessments

Follow-up for breakthrough SARS-CoV-2 infection (primary outcome, sub-study 1) was from 2 weeks after the second dose of SARS-CoV-2 vaccine up to 6 months thereafter or the date of breakthrough SARS-CoV-2 infection, whichever was earlier. Vaccination details and breakthrough SARS-CoV-2 infections confirmed by RT-PCR or antigen testing were captured via online questionnaires sent to all participants at monthly intervals, and complemented by electronic linkage to routinely collected medical record data, as previously described [21]. Every monthly questionnaire contained the following advice to encourage participants with COVID-19 symptoms to engage with testing services: “If you currently have symptoms of coronavirus (a high temperature, a new, continuous cough or loss of or altered sense of smell or taste), call NHS111 or visit https://www.nhs.uk/conditions/coronavirus-covid-19/ for more information.” This wording was identical for questionnaires sent to participants randomised to intervention or no-offer groups. In addition to monthly questionnaires, an online adherence questionnaire was sent to all participants randomised to either offer on 31 March 2021. This questionnaire captured information regarding frequency of study supplement use. End-trial postal vitamin D testing was offered to a randomly selected subset of 1600 participants who received study supplements (800 participants from each intervention group) and 400 who were randomised to no offer. Participants randomised to no offer who were found to have end-trial 25(OH)D concentrations below 50 nmol/L were posted a 60-day supply of capsules each containing 2500 IU vitamin D_3_ (Cytoplan Ltd).

### 2.6. Laboratory assays

#### 2.6.1. 25(OH)D testing

25(OH)D assays were performed by Black Country Pathology Services, located at Sandwell General Hospital, West Bromwich, UK; this laboratory participates in the UK NEQAS for Vitamin D and the Vitamin D External Quality Assessment Scheme (DEQAS) for serum 25(OH)D. Concentrations of 25(OH)D_3_ and 25(OH)D_2_ were determined in dried blood spot eluates using liquid chromatography tandem mass spectrometry (Acquity UPLC-TQS or TQS-Micro Mass Spectrometers, Waters Corp., Milford, MA, USA) after derivatisation and liquid–liquid extraction as previously described [24] and summed to give total 25(OH)D concentrations. Very good overall agreement between blood spot and plasma 25(OH)D concentrations in paired capillary and venous samples using this blood spot method has been observed [24], demonstrating a minimal overall bias of -0.2% with a bias range of -16.9% to 26.7%. Total 25(OH)D concentrations lower than 75 nmol/L were defined as sub-optimal: this threshold is widely considered to discriminate between those with lower vs. higher vitamin D status [25-27]. The between-day coefficients of variation were 11.1% at 16.9 nmol/L, 8.2% at 45.5 nmol/L, 6.9% at 131.7 nmol/L and 7.0% at 222.2 nmol/L for 25(OH)D_3_ and 13.7% at 18.1 nmol/L, 7.5% at 42.7 nmol/L and 6.4% at 127.3 nmol/L for 25(OH)D_2_. The mean bias of dried blood spot vs. serum 25(OH)D_3_ concentrations over the period 2018 to 2021 was 4.0% and the limits of quantitation were 7.5 nmol/L for 25(OH)D_3_ and 2.8 nmol/L for 25(OH)D_2_.

#### 2.6.2. Anti-S serology testing

Anti-S antibody titres were determined by the Clinical Immunology Service at the Institute of Immunology and Immunotherapy of the University of Birmingham (Birmingham, UK) using an ELISA that measures combined IgGAM responses to the SARS-CoV-2 trimeric S glycoprotein (product code MK654; The Binding Site [TBS], Birmingham, UK), as previously described [17]. This assay has been CE-marked with 98.3% (95% confidence interval [CI] 96.4–99.4) specificity and 98.6% (92.6– 100.0) sensitivity for RT-PCR-confirmed mild-to-moderate COVID-19 [28], and has been validated as a correlate of protection against breakthrough SARS-CoV-2 infection in two populations [29, 30]. A cut-off ratio relative to the TBS cut-off calibrators was determined by plotting 624 pre-2019 negatives in a frequency histogram. A cut-off coefficient was then established for IgGAM (1.31), with ratio values classed as positive (≥1) or negative (<1). Dried blood spot eluates were pre-diluted 1:40 with 0.05% PBS-Tween using a Dynex Revelation automated absorbance microplate reader (Dynex Technologies; Chantilly, VA, USA). Plates were developed after 10 min using 3,3′,5,5′-tetramethylbenzidine core, and orthophosphoric acid used as a stop solution (both TBS). Optical densities at 450 nm were measured using the Dynex Revelation.

#### 2.6.3. SARS-CoV-2 neutralising antibody

Serum titres of neutralising antibodies to SARS-CoV-2 were measured as previously described using an authentic virus (Wuhan Hu-1 strain) neutralisation assay [31].

#### 2.6.4. Whole blood stimulation assay

Peripheral blood was collected into heparinised tubes and cultured in the presence or absence of *E*.*coli* lipopolysaccharide (LPS, 1–1000 ng/mL; Invivogen) or PepTivator® SARS-CoV-2 Prot_S Complete (Miltenyi Biotec; 1 µg/mL) for 24 hours at 37°C in 5% CO_2_. Supernatants were harvested and stored at -80°C pending determination of cytokine concentrations by cytometric bead array.

#### 2.6.5. Cytometric bead array

Cytometric bead array of whole blood assay supernatants was carried out according to the manufacturer’s protocol (BD Biosciences). The cytokines assessed were IL-6, IL-8, IFN-γ and TNF. Samples were analysed using the ACEA Novocyte 3000 flow cytometer (Agilent). The lower limit of detection was 1.5 pg/mL.

#### 2.6.6. Peripheral blood mononuclear cell (PBMC) isolation

PBMC were isolated from heparinised blood using Ficoll (Merck Life Science) density gradient, washed twice in Hanks Balanced Salt Solution (Merck Life Science) and cryopreserved in 10% DMSO in Fetal Calf Serum (Invitrogen).

#### 2.6.7. PBMC stimulation assay

Cryopreserved PBMC were recovered and stimulated with nothing (negative control) or PepTivator® SARS-CoV-2 Prot_S Complete (Miltenyi Biotec; 1 µg/mL) or 1 µg/mL of soluble CD3 Monoclonal Antibody (OKT3), Functional Grade (Invitrogen) for 1 hour at 37°C in a 5% CO_2_. Brefeldin A (2.5 µg/mL) was then added to the cells, which were incubated for a further 15 hours at 37°C in 5% CO_2_.

#### 2.6.8. Flow cytometric analysis

Stimulated cells were collected, and cell surface stained for CD3 (HIT3a), CD4 (RPA-T4), CD8 (SK1), CD27 (O232), CD45RA (HI100) and Zombie NIR™ viability dye (Biolegend) in the presence of Brilliant Buffer (BD Biosciences). Cells were washed and fixed in Intracellular Fixation Buffer (eBioscience), permeabilised in eBioscience Permeablization Buffer, stained for intracellular IL-2 (JES6-5H4), IFN-γ (4S.B3) and TNF (Mab11, Biolegend), washed and assessed using the ACEA Novocyte 3000 flow cytometer (Agilent). Data were analysed using FlowJo Version X (BD Biosciences).

### 2.7. Outcomes

Outcomes for sub-study 1 were time to breakthrough SARS-CoV-2 infection, and the proportion of participants experiencing an episode of breakthrough infection during follow-up. Follow-up for this efficacy analysis began 14 days after participants received a second vaccine dose and participants were censored either at time of breakthrough infection, 13 days after their booster dose, or at 6 months’ follow-up, whichever occurred earlier. Outcomes for sub-study 2 were anti-S titres and the proportion of participants with detectable anti-S antibodies after vaccination. Outcomes for sub-study 3 were neutralising antibody titres; concentrations of IFN-γ, TNF, IL-6 and CXCL8 in supernatants of S peptide- and LPS-stimulated whole blood; percentages of S-peptide and CD3-stimulated T cell subsets staining positive for IFN-γ, IL-2 and TNF; and percentages of T cell subsets with naÏve, central memory, effector memory and terminally differentiated effector memory cells re-expressing CD45RA (EMRA) phenotypes after vaccination.

### 2.8. Statistical methods

Trial sample size was calculated using https://mjgrayling.shinyapps.io/multiarm/ [32], and predicated on numbers needed to detect a 20% reduction in the proportion of participants experiencing one or more acute respiratory infections with 84% marginal power and 5% type 1 error rate, as described elsewhere [21].

Statistical analyses were performed using Stata version 17.0. Pairwise comparisons were made between each intervention arm separately vs. the no-offer arm, and between pooled data from participants randomised to either intervention arm vs. the no-offer arm. Time to breakthrough SARS-CoV-2 infections was compared between study arms using Cox regression, with adjustment for factors we have previously reported to be risk factors for breakthrough SARS-CoV-2 infection: age, sex, educational attainment, frontline worker status, number of people per bedroom, sharing a home with schoolchildren (5–15 years), primary vaccination course, previous SARS-CoV-2 infection, season of first vaccination, inter-dose interval, use of anticholinergics, weekly visits to or from other households, weekly visits to indoor public places other than shops, and local weekly SARS-CoV-2 incidence (according to participants’ area of residence). Treatment effects are presented as adjusted hazard ratios (aHRs) with 95% CIs. Proportions of participants experiencing breakthrough SARS-CoV-2 infection were compared between arms using logistic regression, with adjustment for the same covariates and presentation of treatment effects as adjusted odds ratios (aORs) with 95% CIs. Linear regression was used to estimate inter-arm geometric mean ratios (GMRs), with 95% CIs and associated pairwise *p*-values, for log-transformed antibody titres, cytokine concentrations, and percentages of T cell subsets staining positive for intracellular cytokines and exhibiting different phenotypes, with adjustment for factors we have previously shown to be determinants of post-vaccination anti-S titres [17]: age, sex, ethnicity, body-mass index, days from second vaccine dose to DBS sample, pre-vaccination serostatus, general health, inter-dose interval and primary vaccination course. For ease of interpretation, estimated GMRs are expressed as adjusted percentage differences. For immunological outcomes, correction for multiple comparisons was performed on families of pairwise *p*-values using the Benjamini & Hochberg method with a false discovery rate of 5% [33].

We conducted a sensitivity analysis for outcomes of breakthrough SARS-CoV-2 infection, anti-S titre and S peptide-stimulated IFN-γ, excluding data from participants who reported a SARS-CoV-2 infection prior to vaccination.

## 3. Results

### 3.1. Participants

Of 6200 CORONAVIT trial participants, 2808 (45.3%) received a primary course of SARS-CoV-2 vaccination (with first doses administered between 16 January and 16 June 2021) and contributed data to sub-study 1 (vaccine efficacy analysis); of these, 1945 (69.3%) received two doses of ChAdOx1 nCoV-19 and 863 (30.7%) received two doses of BNT162b2. 1853 (30.0%) provided a post-vaccination dried blood spot sample between 22 March 2021 and 16 November 2021 and contributed data to sub-study 2 (analysis of anti-S IgGAM titres) and 101 (1.6%) provided a post-vaccination venous blood sample between 24 May 2021 and 12 August 2021 and contributed data to the analysis of neutralising antibody and cellular responses to SARS-CoV-2 vaccination (sub-study 3; Fig. 1). Table 1 shows baseline characteristics of participants included in the vaccine efficacy analysis (sub-study 1) by allocation. Median age was 61.9 years, 65.8% were female, and 96.4% were of White ethnic origin. Among participants whose baseline vitamin D status was tested, mean 25(OH)D concentration was 39.9 nmol/L, and all had 25(OH)D concentrations below 75 nmol/L. Characteristics were balanced between the three trial arms, except for proportions of participants with pre-vaccination SARS-CoV-2 infection (4.2 vs. 5.9 vs. 2.9% in no offer vs. 800 IU/day vs. 3200 IU/day arms, respectively). Baseline characteristics for participants additionally contributing data to analyses of anti-S titres (sub-study 2) and neutralising antibody or cellular responses (sub-study 3) are presented in Tables S1 and S2 of the Supplementary Appendix, respectively: these were also balanced between trial arms. Among participants for whom end-study vitamin D measurements were available, mean follow-up 25(OH)D concentrations were significantly elevated in the lower-dose vs. no-offer group (82.5 [standard deviation 18.9] vs. 53.6 [25.2] nmol/L; mean difference 28.8 nmol/L, 95% CI 22.8– 34.8) and in the higher-dose vs. no offer group (105.4 [23.5] vs. 53.6 [25.2] nmol/L; mean difference 51.7 nmol/L, 45.1–58.4; Fig. 2A).

**Figure 1:**
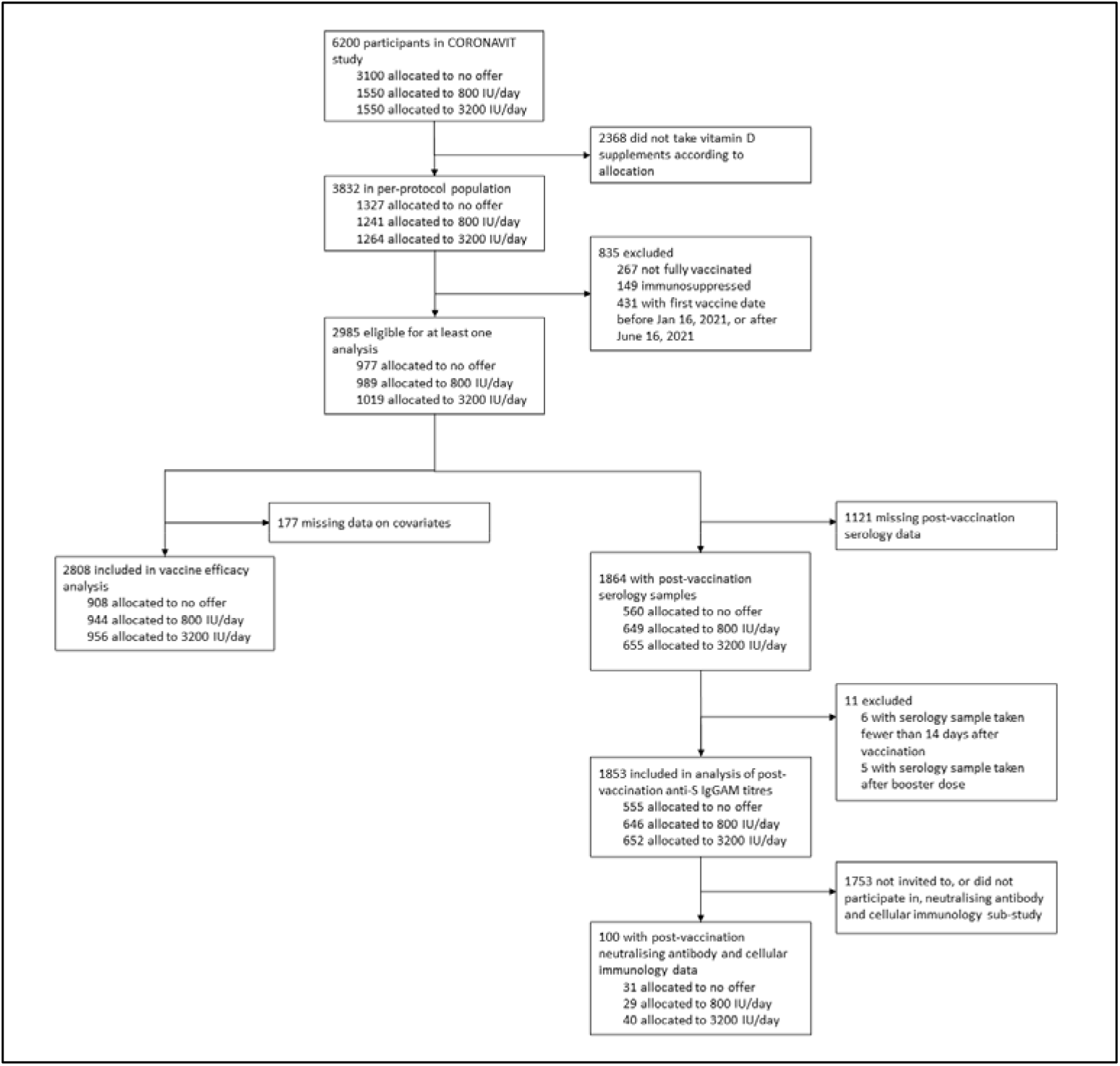
Participant flow.

**Figure 2:**
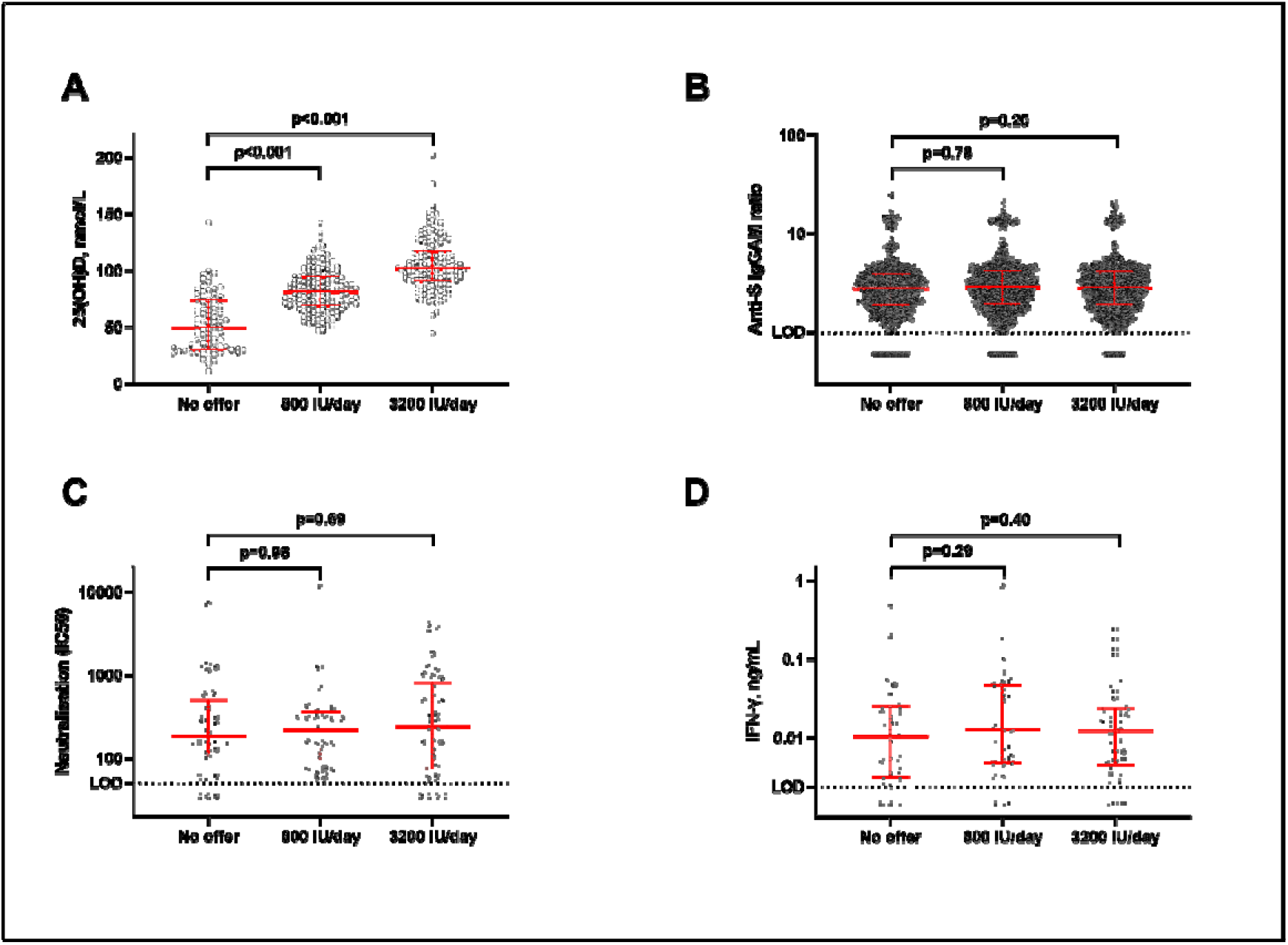
Biochemical and immunological outcomes by allocation. (A) End-study 25(OH)D concentrations. (B) Anti-S IgGAM titres. (C) Neutralising antibody titres. (D) IFN-_γ_ concentrations in supernatants from S peptide-stimulated whole blood. Horizontal bars represent medians and interquartile ranges. P values from unpaired *t* tests (A) and multiple linear regression with adjustment for covariates as described in Methods (B–D). 25(OH)D=25-hydroxyvitamin D. Anti-S IgGAM=combined anti-Spike IgG, IgA and IgM response. IFN=interferon. LOD=limit of detection.

**Table 1.** Baseline characteristics of participants contributing data to vaccine efficacy analysis (sub-study 1), by allocation.

|  | Overall (n=2808) | No offer (n=908) | 800 IU/day offer (n=944) | 3200 IU/day offer (n=956) |
| --- | --- | --- | --- | --- |
| Age, years |  |  |  |  |
| Median (IQR) | 61.9 (54.1–68.5) | 61.8 (53.6–68.1) | 61.4 (54.2–68.3) | 62.5 (54.1–69.1) |
| Range | 16.6–87.2 | 18.4–85.9 | 20.9–83.1 | 16.6–87.2 |
| Sex |  |  |  |  |
| Female | 1,848 (65.8%) | 579 (63.8%) | 634 (67.2%) | 635 (66.4%) |
| Male | 960 (34.2%) | 329 (36.2%) | 310 (32.8%) | 321 (33.6%) |
| Ethnicity |  |  |  |  |
| White | 2,706 (96.4%) | 885 (97.5%) | 910 (96.4%) | 911 (95.3%) |
| Asian/Asian British | 25 (0.9%) | 9 (1.0%) | 5 (0.5%) | 11 (1.2%) |
| Black/African/Caribbean/Black British | 14 (0.5%) | 2 (0.2%) | 6 (0.6%) | 6 (0.6%) |
| Mixed/Multiple/Other | 63 (2.2%) | 12 (1.3%) | 23 (2.4%) | 28 (2.9%) |
| Body-mass index, kg/m<sup>2</sup> |  |  |  |  |
| <25 | 1,304 (46.5%) | 404 (44.5%) | 440 (46.8%) | 460 (48.1%) |
| 25–30 | 949 (33.8%) | 314 (34.6%) | 307 (32.6%) | 328 (34.3%) |
| >30 | 551 (19.7%) | 189 (20.8%) | 194 (20.6%) | 168 (17.6%) |
| Self-assessed general health |  |  |  |  |
| Excellent | 617 (22.0%) | 203 (22.4%) | 196 (20.8%) | 218 (22.8%) |
| Very good | 1,186 (42.3%) | 386 (42.5%) | 404 (42.8%) | 396 (41.5%) |
| Good | 721 (25.7%) | 213 (23.5%) | 257 (27.2%) | 251 (26.3%) |
| Fair | 222 (7.9%) | 86 (9.5%) | 67 (7.1%) | 69 (7.2%) |
| Poor | 61 (2.2%) | 20 (2.2%) | 20 (2.1%) | 21 (2.2%) |
| Medically diagnosed disease |  |  |  |  |
| Hypertension | 577 (20.5%) | 191 (21.0%) | 190 (20.1%) | 196 (20.5%) |
| Diabetes | 135 (4.8%) | 50 (5.5%) | 35 (3.7%) | 50 (5.2%) |
| Heart disease | 113 (4.0%) | 41 (4.5%) | 39 (4.1%) | 33 (3.5%) |
| Asthma | 394 (14.0%) | 120 (13.2%) | 160 (16.9%) | 114 (11.9%) |
| COPD | 45 (1.6%) | 18 (2.0%) | 15 (1.6%) | 12 (1.3%) |
| Pre-vaccination SARS-CoV-2 infection<sup>(1)</sup> | 121 (4.3%) | 38 (4.2%) | 55 (5.8%) | 28 (2.9%) |
| Type of vaccine administered, primary course<sup>(2)</sup> |  |  |  |  |
| 2 × ChAdOx1 | 1,945 (69.3%) | 632 (69.6%) | 673 (71.3%) | 640 (66.9%) |
| 2 × BNT162b2 | 863 (30.7%) | 276 (30.4%) | 271 (28.7%) | 316 (33.1%) |
| Inter-dose interval, days | 77 (69–79) | 77 (68–79) | 77 (69–79) | 77 (69–79) |
| Mean 25(OH)D, nmol/L (SD) [range]<sup>(3)</sup> | 39.9 (14.5) [10.3–74.9] | --<sup>(4)</sup> | 39.6 (14.7) [10.3–74.8] | 40.2 (14.4) [10.3–74.9] |
| <25.0 | 309 (11.0%) | --<sup>(4)</sup> | 164 (17.4%) | 145 (15.2%) |
| 25.0 to <50.0 | 1,104 (39.3%) | --<sup>(4)</sup> | 543 (57.5%) | 561 (58.7%) |
| 50.0 to <75.0 | 482 (17.2%) | --<sup>(4)</sup> | 235 (24.9%) | 247 (25.8%) |
| ≥75.0 | 0 | --<sup>(4)</sup> | 0 | 0 |
| Not determined | 913 (32.5%) | 908 (100.0%) | 2 (0.2%) | 3 (0.3%) |

Data are n (%) or median (IQR) unless specified otherwise. Abbreviations: IQR, interquartile range; SD, standard deviation; 25(OH)D, 25-hydroxyvitamin D. (1) Reported swab test-positive infection by RT-PCR or antigen testing. (2) Vaccine efficacy analysis restricted to participants receiving a full course of either ChAdOx1 or BNT162b2. (3) Missing values: 25(OH)D concentration missing for three participants in 3200 IU/day arm and two participants in 800 IU/day arm. (4) Baseline 25(OH)D not determined for participants randomly assigned to the no-offer arm.

**Table 2.**
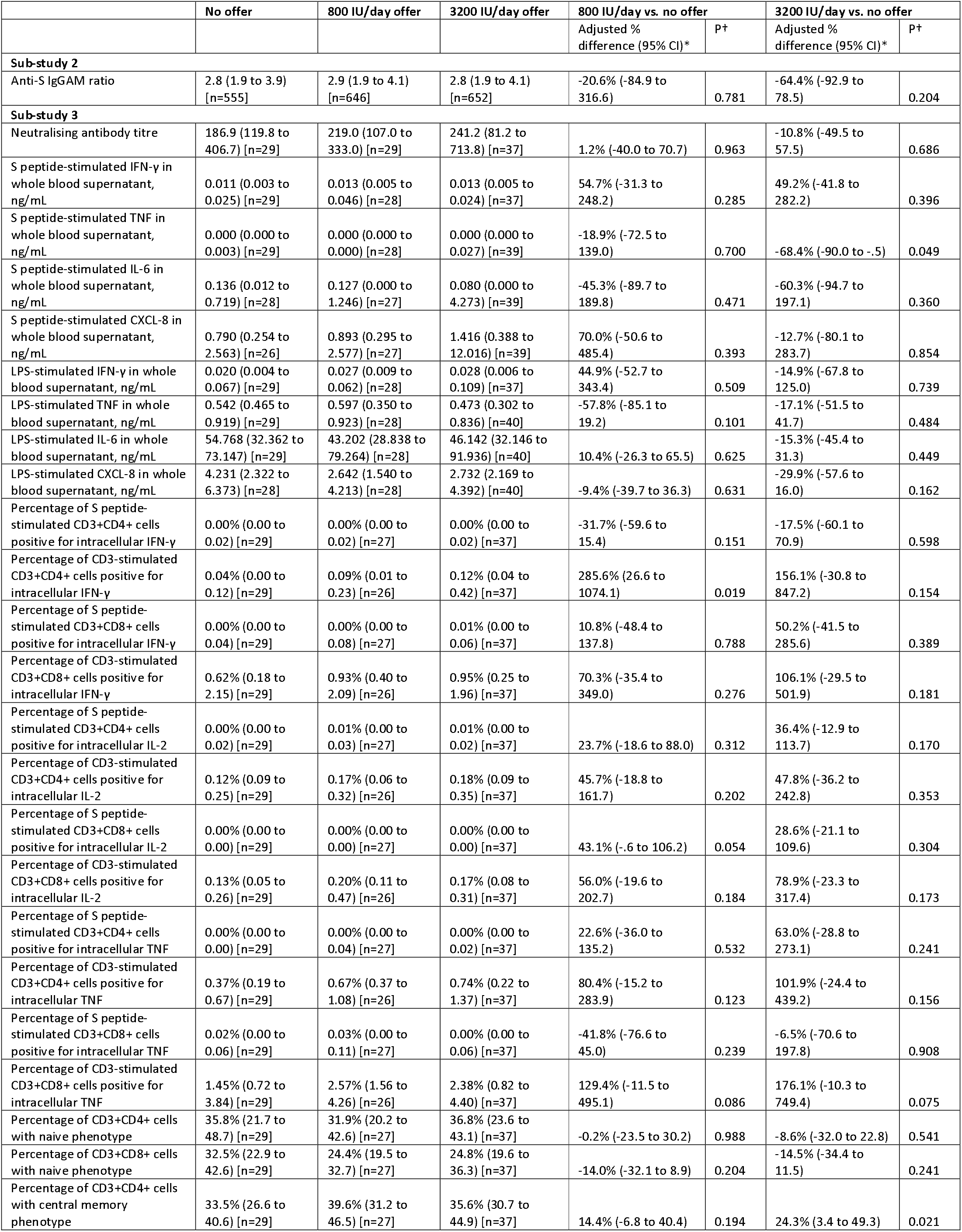
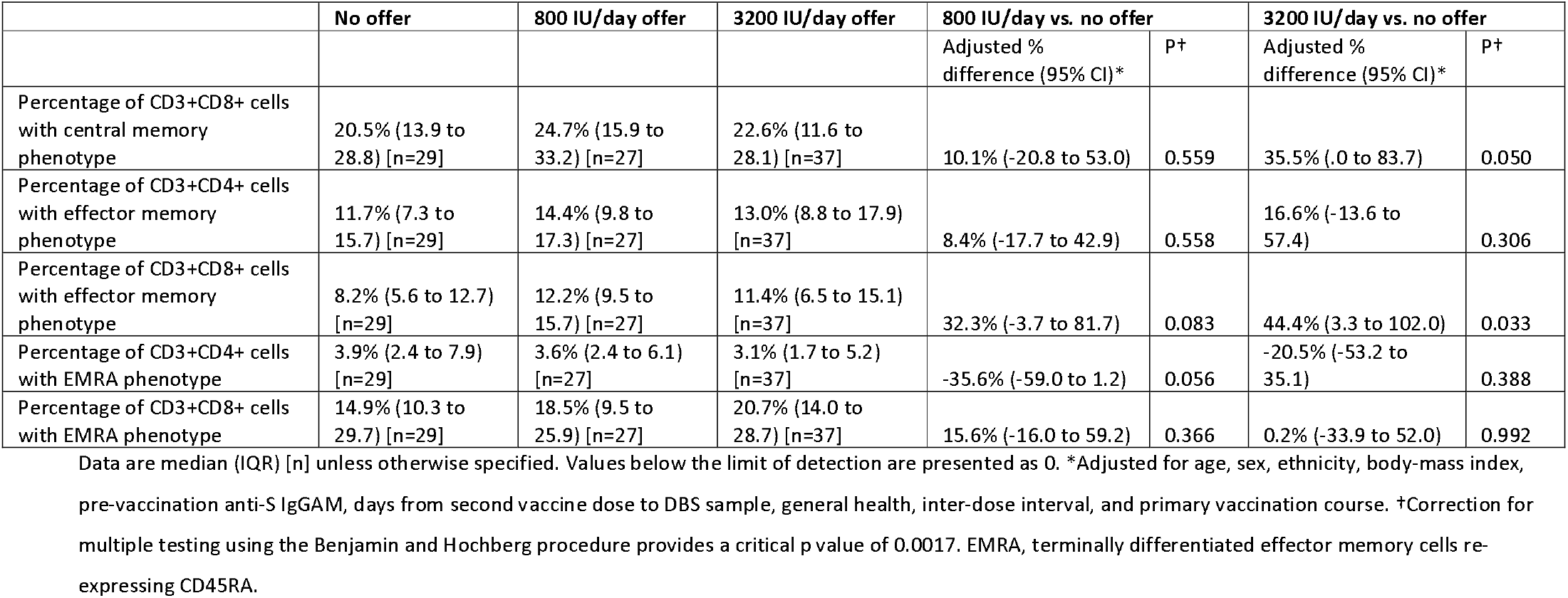
Immunological outcomes by allocation.

### 3.2. Breakthrough SARS-CoV-2 infection

Breakthrough SARS-CoV-2 infection occurred in 174 sub-study 1 participants, with no significant inter-arm difference in time to event (lower dose vs. no offer: median 160 [IQR 114–192] vs. 161 [131–193] days to infection, aHR 1.28, 95% CI 0.89– 1.84, *p*□=□0.19; higher dose vs. no offer: 146 [88–189] vs. 160 [131–193] days to infection, aHR 1.17, 0.81–1.70, *p*□=□0.40; Fig. 3A, 3B). Results were similar when pooling data from both intervention arms (any offer vs. no offer: 153 [106–192] vs. 161 [131–193] days to infection, aHR 1.24, 0.89–1.71, *p*□=□0.20; Fig. 3C). Consistent with these findings, proportions of participants experiencing breakthrough infection did not differ by allocation (Table S3, Supplementary Appendix).

**Figure 3:**
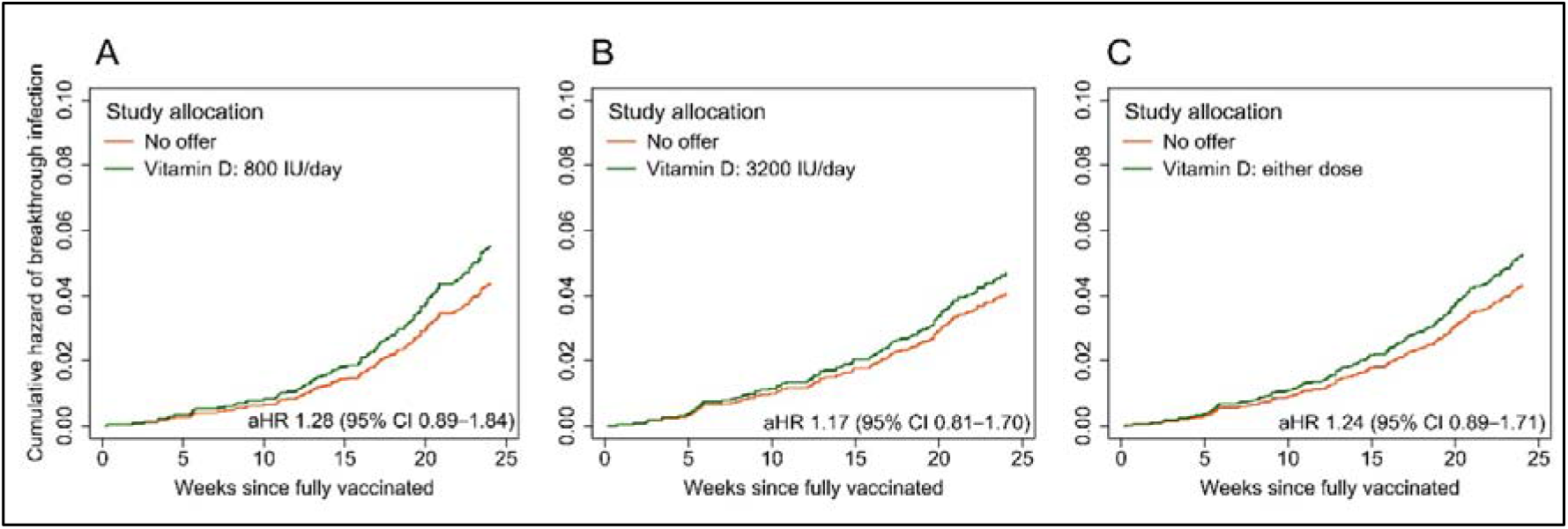
Cumulative hazard plots showing risk of breakthrough SARS-CoV-2 infection by allocation. A, offer of 800 IU vitamin D3/day vs. no offer. B, offer of 3200 IU vitamin D3/day vs. no offer. C, either offer (pooled) vs. no offer. Hazard ratios adjusted for age, sex, educational attainment, frontline worker status, number of people per bedroom, schoolchildren (5–15 years) at home with participant, primary vaccination course, previous SARS-CoV-2 infection, season of first vaccination, inter-dose interval, use of anticholinergics, weekly visits to or from other households, weekly visits to indoor public places other than shops, and local weekly SARS-CoV-2 incidence in participants’ area of residence. aHR, adjusted hazard ratio.

### 3.3. Immunological outcomes

No inter-arm differences in mean post-vaccination titres of combined anti-S IgGAM antibodies were seen, either when each intervention arm was compared separately to the no-offer arm (Table 3, Fig. 2B) or when pooled data from both intervention arms were compared to the no-offer arm (Table S4, Supplementary Appendix). Neither was there any inter-arm difference in proportions of participants with detectable post-vaccination anti-S IgGAM antibodies (Table S3, Supplementary Appendix). In the subset of participants who provided a venous blood sample for analysis, we found no significant inter-arm differences in mean neutralising antibody titres or in any antigen-specific cellular response investigated after correction for multiple testing, either when lower-dose or higher-dose arms were compared separately to the no-offer arm (Table 3, Fig. 2C, 2D) or when pooled data from both intervention arms were compared to the no-offer arm (Table S4, Supplementary Appendix).

### 3.4. Sensitivity analysis

Excluding participants with previous SARS-CoV-2 infection did not substantially affect our findings on breakthrough SARS-CoV-2 infection (Tables S5, 26, Supplementary Appendix), anti-S IgGAM titres (Tables S6, S7, Supplementary Appendix), or S peptide-stimulated IFN-γ (Table S7, Supplementary Appendix).

## 4. Discussion

We report findings of sub-studies nested within a randomised controlled trial to investigate effects of vitamin D supplementation on SARS-CoV-2 vaccine efficacy and immunogenicity. All participants had 25(OH)D concentrations below 75 nmol/L at baseline, and supplementation with both 800 IU and 3200 IU of vitamin D per day was effective in elevating end-study 25(OH)D concentrations in the intervention groups. However, improvements in vitamin D status were not associated with inter-arm differences in risk of breakthrough SARS-CoV-2 infection, post-vaccination titres of anti-S or neutralising antibodies or any cellular immune response investigated.

Null results from the current intervention study are in keeping with those from two observational studies in the field [19, 20], but contrast with findings from two others that report positive results. Of these, one reported an association between higher post-vaccination anti-S titres and circulating 25(OH)D concentrations of more than 50 nmol/L in a cohort of health care workers [18]. The other, a population-based study conducted in UK adults, found an independent association between vitamin D supplement use and reduced risk of anti-S seronegativity following SARS-CoV-2 vaccination [17]. These positive associations may have arisen as a result of unmeasured or residual confounding, or type 1 error. The fact that no inter-arm difference in anti-S titres was seen in the current study supports the interpretation that the null result from the current analysis is valid, since it is biologically implausible that vitamin D would affect the proportion of seronegative participants but not the mean anti-S titre. The null findings presented here also contrast with results of our previous intervention study [14], in which we showed that vitamin D replacement in older adults with baseline 25(OH)D levels below 75 nmol/L boosted antigen-specific immunity and reduced inflammatory responses to cutaneous VZV antigen challenge. Divergent findings between these two intervention studies may reflect differences in the compartment studied (peripheral blood vs. skin), immunological stimulus (SARS-CoV-2 vaccination vs. VZV antigen challenge) or the regimen of vitamin D administered (800 or 3200 IU/day for at least 1 month vs. 6400 IU/day for 14 weeks before stimulation).

Our study has several strengths. Participants had sub-optimal vitamin D status at baseline, and interventions were effective in elevating 25(OH)D levels into the physiological range. The large sample sizes of sub-studies 1 and 2, together with the substantial number of breakthrough SARS-CoV-2 infections arising in sub-study 1, provided good power to detect effects of the intervention. We also investigated a combination of clinical and immunological outcomes, with detailed characterisation of both humoral and cellular responses: the fact that our results were consistent across a broad range of outcomes strengthens the interpretation that our null results are valid.

Our study also has limitations. Randomisation could not be stratified according to sub-study participation, since eligibility for inclusion in one or more sub-studies was contingent on factors arising during follow-up. However, baseline characteristics were similar between arms for all sub-studies, and we adjusted for multiple factors influencing vaccine efficacy and immunogenicity, thereby minimising the potential for confounding. There was a baseline imbalance in pre-vaccination SARS-CoV-2 status between study arms. We accounted for this in the analysis by adjustment in the main model, and by conducting a sensitivity analysis excluding data from participants who had pre-vaccination SARS-CoV-2 infection at baseline: these complementary approaches yielded similar results. Our study was open label, and participants were therefore aware of their allocation: however, laboratory staff were blinded to participant allocation, thereby removing the potential for observer bias to influence assessment of immunological outcomes. No restrictions regarding vitamin D intake were stipulated for participants randomised to the no-offer arm; however, participants in this group who reported taking supplements were excluded from the analyses presented here. The fact that attained 25(OH)D concentrations differed markedly between arms suggests that we were successful in excluding participants in the no-offer arm who used off-trial vitamin D supplements during follow-up. Finally, the lack of a measurement of baseline vitamin D status among participants in the no-offer arm precludes sub-group analyses to test for this as an effect modifier. Although all participants tested had baseline 25(OH)D concentrations below 75 nmol/L, we cannot rule out an effect in sub-groups of participants with the lowest baseline 25(OH)D concentrations.

In conclusion, we report that daily administration of 800 IU or 3200 IU vitamin D3 was effective in elevating circulating 25(OH)D concentrations, but that neither dose influenced SARS-CoV-2 vaccine efficacy or immunogenicity. Our findings do not support the use of vitamin D supplements as an adjunct to SARS-CoV-2 vaccination.

## Supporting information

Supplementary Appendix Jolliffe et al

## Data Availability

Anonymised participant-level data will be made available on reasonable request to, subject to the terms of Research Ethics Committee and Sponsor approval.

## Contributors

ARM, DAJ and ESC designed the sub-studies. The trial was managed by DAJ and ARM. Laboratory assays were developed and/or performed by ESC, WC, WL, SEF, JMG, CP, AGR and AMcK. Data were analysed by GV. ARM wrote the first draft of the paper. All authors contributed to the interpretation of the results, review and approval of the manuscript, and the decision to submit it for publication.

## Ethical approval

The study was approved by the Queens Square Research Ethics Committee, London, UK (ref 20/HRA/5095) and all participants gave informed consent before taking part.

## Transparency statement

ARM affirms that the manuscript is an honest, accurate, and transparent account of the study results; that no important aspects of the study have been omitted; and that any discrepancies from the study as planned and registered have been explained. All other outcomes pre-specified in the protocol and the trial registry have been reported elsewhere (https://doi.org/10.1101/2022.03.22.22271707).

## Acknowledgements

We thank all the people who participated in the trial; members of the Independent Data Monitoring Committee (Prof Irwin Nazareth, University College London [Chair]; Dr Michael Grayling; and Dr Richard Quinton, University of Newcastle upon Tyne); and members of the Trial Steering Committee (Prof Paul Lips, Amsterdam University Medical Centre, Amsterdam [Chair]; Dr Gwyneth Davies, University College London; and Dr Anna Bibby, University of Bristol).

## References

1 Feikin DR, Higdon MM, Abu-Raddad LJ, et al. Duration of effectiveness of vaccines against SARS-CoV-2 infection and COVID-19 disease: results of a systematic review and meta-regression. Lancet 2022; 399: 924–44.

2 Liang Z, Zhu H, Wang X, et al. Adjuvants for Coronavirus Vaccines. Front Immunol 2020; 11: 589833.

3 Ciabattini A, Nardini C, Santoro F, Garagnani P, Franceschi C, Medaglini D. Vaccination in the elderly: The challenge of immune changes with aging. Seminars in immunology 2018; 40: 83–94.

4 De Maeyer RPH, Chambers ES. The impact of ageing on monocytes and macrophages. Immunol Lett 2020.

5 Vukmanovic-Stejic M, Chambers ES, Suarez-Farinas M, et al. Enhancement of cutaneous immunity during aging by blocking p38 mitogen-activated protein (MAP) kinase-induced inflammation. J Allergy Clin Immunol 2018; 142: 844–56.

6 Mannick JB, Del Giudice G, Lattanzi M, et al. mTOR inhibition improves immune function in the elderly. Sci Transl Med 2014; 6: 268ra179.

7 Mannick JB, Morris M, Hockey HP, et al. TORC1 inhibition enhances immune function and reduces infections in the elderly. Sci Transl Med 2018; 10.

8 Bikle DD. Vitamin D Regulation of Immune Function. Current osteoporosis reports 2022; 20: 186–93.

9 Zhang Y, Leung DY, Richers BN, Liu Y, Remigio LK, Riches DW, Goleva E. Vitamin D inhibits monocyte/macrophage proinflammatory cytokine production by targeting MAPK phosphatase-1. J Immunol 2012; 188: 2127–35.

10 Lisse TS, Hewison M. Vitamin D: a new player in the world of mTOR signaling. Cell Cycle 2011; 10: 1888–9.

11 von Essen MR, Kongsbak M, Schjerling P, Olgaard K, Odum N, Geisler C. Vitamin D controls T cell antigen receptor signaling and activation of human T cells. Nature immunology 2010; 11: 344–9.

12 De Vita F, Lauretani F, Bauer J, et al. Relationship between vitamin D and inflammatory markers in older individuals. Age (Dordr) 2014; 36: 9694.

13 Laird E, McNulty H, Ward M, et al. Vitamin D deficiency is associated with inflammation in older Irish adults. J Clin Endocrinol Metab 2014; 99: 1807–15.

14 Chambers ES, Vukmanovic-Stejic M, Turner CT, et al. Vitamin D3 replacement enhances antigen-specific immunity in older adults. Immunother Adv 2020.

15 Calder PC, Berger MM, Gombart AF, McComsey GA, Martineau AR, Eggersdorfer M. Micronutrients to Support Vaccine Immunogenicity and Efficacy. Vaccines (Basel) 2022; 10.

16 Chiu SK, Tsai KW, Wu CC, et al. Putative Role of Vitamin D for COVID-19 Vaccination. Int J Mol Sci 2021; 22.

17 Jolliffe DA, Faustini SE, Holt H, et al. Determinants of Antibody Responses to Two Doses of ChAdOx1 nCoV-19 or Bnt162b2 and a Subsequent Booster Dose of BNT162b2 or mRNA-1273: Population-Based Cohort Study (COVIDENCE UK). Preprints with The Lancet, 2022.

18 Piec I, Cook L, Dervisevic S, et al. Age and vitamin D affect the magnitude of the antibody response to the first dose of the SARS-CoV-2 BNT162b2 vaccine. Curr Res Transl Med 2022; 70: 103344.

19 Chillon TS, Demircan K, Heller RA, et al. Relationship between Vitamin D Status and Antibody Response to COVID-19 mRNA Vaccination in Healthy Adults. Biomedicines 2021; 9.

20 Parthymou A, Habeos EE, Habeos GI, Deligakis A, Livieratos E, Marangos M, Chartoumpekis DV. Factors associated with anti-SARS-CoV-2 antibody titres 3 months post-vaccination with the second dose of BNT162b2 vaccine: a longitudinal observational cohort study in western Greece. BMJ Open 2022; 12: e057084.

21 Jolliffe DA, Holt H, Greenig M, et al. Vitamin D Supplements for Prevention of Covid-19 or other Acute Respiratory Infections: a Phase 3 Randomized Controlled Trial (CORONAVIT). MedRxiv 2022.

22 Hypponen E, Power C. Hypovitaminosis D in British adults at age 45 y: nationwide cohort study of dietary and lifestyle predictors. Am J Clin Nutr 2007; 85: 860–8.

23 Holt H, Relton C, Talaei M, et al. Cohort Profile: Longitudinal population-based study of COVID-19 in UK adults (COVIDENCE UK). MedRxiv 2022.

24 Shea RL, Berg JD. Self-administration of vitamin D supplements in the general public may be associated with high 25-hydroxyvitamin D concentrations. Ann Clin Biochem 2017; 54: 355–61.

25 Dawson-Hughes B, Heaney RP, Holick MF, Lips P, Meunier PJ, Vieth R. Estimates of optimal vitamin D status. Osteoporos Int 2005; 16: 713–6.

26 Vieth R. What is the optimal vitamin D status for health? Prog Biophys Mol Biol 2006; 92: 26–32.

27 Bischoff-Ferrari HA. The 25-hydroxyvitamin D threshold for better health. J Steroid Biochem Mol Biol 2007; 103: 614–9.

28 Cook AM, Faustini SE, Williams LJ, et al. Validation of a combined ELISA to detect IgG, IgA and IgM antibody responses to SARS-CoV-2 in mild or moderate non-hospitalised patients. Journal of Immunological Methods 2021; 494: 113046.

29 Shields AM, Faustini SE, Kristunas CA, et al. COVID-19: Seroprevalence and Vaccine Responses in UK Dental Care Professionals. J Dent Res 2021; 100: 1220–7.

30 Vivaldi G, Jolliffe DA, Faustini SE, et al. Correlation between post-vaccination titres of IgG, IgA and IgM anti-Spike antibodies and protection against breakthrough SARS-CoV-2 infection: a population-based longitudinal study (COVIDENCE UK). MedRxiv 2022.

31 Reynolds CJ, Swadling L, Gibbons JM, et al. Discordant neutralizing antibody and T cell responses in asymptomatic and mild SARS-CoV-2 infection. Sci Immunol 2020; 5.

32 Grayling MJ, Wason JM. A web application for the design of multi-arm clinical trials. BMC Cancer 2020; 20: 80.

33 Benjamini Y, Hochberg Y. Controlling the False Discovery Rate - a Practical and Powerful Approach to Multiple Testing. J Roy Stat Soc B Met 1995; 57: 289–300.

