## Supplementary Appendix Jolliffe et al for "Influence of vitamin D supplementation on SARS-CoV-2 vaccine efficacy and immunogenicity"

### **Table S1.** Baseline characteristics of participants contributing data to analysis of post-vaccination anti-S IgGAM titres (sub-study 2), by allocation

|  | **Overall (n=1853)** | **No offer (n=555)** | **800 IU/day offer (n=646)** | **3200 IU/day offer (n=652)** |
| --- | --- | --- | --- | --- |
| Age, years |  |  |  |  |
| Median (IQR) | 63.6 (57.0–69.3) | 63.5 (57.2–69.2) | 63.0 (56.5–68.8) | 64.1 (57.0–70.1) |
| Range | 16.6–87.2 | 18.4–82.8 | 26.8–83.1 | 16.6–87.2 |
| Sex |  |  |  |  |
| Female | 1,228 (66.3%) | 341 (61.4%) | 447 (69.2%) | 440 (67.5%) |
| Male | 625 (33.7%) | 214 (38.6%) | 199 (30.8%) | 212 (32.5%) |
| Ethnicity |  |  |  |  |
| White | 1,805 (97.4%) | 546 (98.4%) | 629 (97.4%) | 630 (96.6%) |
| Asian/Asian British | 12 (0.6%) | 4 (0.7%) | 0 | 8 (1.2%) |
| Black/African/Caribbean/Black British | 4 (0.2%) | 1 (0.2%) | 2 (0.3%) | 1 (0.2%) |
| Mixed/Multiple/Other | 32 (1.7%) | 4 (0.7%) | 15 (2.3%) | 13 (2.0%) |
| Body-mass index, kg/m² |  |  |  |  |
| <25 | 832 (45.0%) | 238 (43.0%) | 289 (44.9%) | 305 (46.8%) |
| 25–30 | 657 (35.5%) | 208 (37.6%) | 216 (33.5%) | 233 (35.7%) |
| >30 | 360 (19.5%) | 107 (19.3%) | 139 (21.6%) | 114 (17.5%) |
| Self-assessed general health |  |  |  |  |
| Excellent | 417 (22.5%) | 131 (23.6%) | 136 (21.1%) | 150 (23.0%) |
| Very good | 786 (42.4%) | 250 (45.0%) | 276 (42.7%) | 260 (39.9%) |
| Good | 469 (25.3%) | 116 (20.9%) | 177 (27.4%) | 176 (27.0%) |
| Fair | 143 (7.7%) | 48 (8.6%) | 45 (7.0%) | 50 (7.7%) |
| Poor | 37 (2.0%) | 10 (1.8%) | 12 (1.9%) | 15 (2.3%) |
| Medically diagnosed disease |  |  |  |  |
| Hypertension | 409 (22.1%) | 131 (23.6%) | 137 (21.2%) | 141 (21.6%) |
| Diabetes | 96 (5.2%) | 33 (5.9%) | 24 (3.7%) | 39 (6.0%) |
| Heart disease | 77 (4.2%) | 25 (4.5%) | 26 (4.0%) | 26 (4.0%) |
| Asthma | 244 (13.2%) | 66 (11.9%) | 102 (15.8%) | 76 (11.7%) |
| COPD | 32 (1.7%) | 12 (2.2%) | 11 (1.7%) | 9 (1.4%) |
| Previous SARS-CoV-2 infection^(1)^ | 75 (4.1%) | 19 (3.5%) | 35 (5.4%) | 21 (3.3%) |
| Type of vaccine administered, primary course |  |  |  |  |
| 2 × ChAdOx1 | 1,280 (69.1%) | 389 (70.1%) | 463 (71.7%) | 428 (65.6%) |
| 2 × BNT162b2 | 559 (30.2%) | 161 (29.0%) | 180 (27.9%) | 218 (33.4%) |
| Other^(2)^ | 14 (0.8%) | 5 (0.9%) | 3 (0.5%) | 6 (0.9%) |
| Inter-dose interval, days | 77 (70–79) | 77 (70–79) | 77 (70–79) | 77 (70–79) |
| Interval from second vaccine dose to DBS sample, days | 54 (41–70) | 56 (41–71) | 52 (41–67) | 55 (41–71) |
| Mean 25(OH)D, nmol/L (SD) [range]^(3)^ | 40.5 (14.3) [10.3–73.3] | --^(4)^ | 40.4 (14.5) [10.3–73.5] | 40.7 (14.2) [10.3–73.2] |
| <25.0 | 183 (9.9%) | --^(4)^ | 96 (14.9%) | 87 (13.3%) |
| 25.0 to <50.0 | 766 (41.3%) | --^(4)^ | 376 (58.2%) | 390 (59.8%) |
| 50.0 to <75.0 | 344 (18.6%) | --^(4)^ | 172 (26.6%) | 172 (26.4%) |
| ≥75.0 | 0 | --^(4)^ | 0 | 0 |
| Not determined | 560 (30.2%) | 555 (100.0%) | 2 (0.3%) | 3 (0.5%) |

Data are n (%) or median (IQR) unless specified otherwise. Abbreviations: anti-S IgGAM, combined anti-Spike IgG, IgA and IgM response; IQR, interquartile range; DBS, dried blood spot; SD, standard deviation; 25(OH)D, 25-hydroxyvitamin D. (1) Reported test-positive infection before provision of DBS sample. (2) One participant receiving 2 × Moderna (3200 IU/day offer), two participants receiving 2 × Novavax (no offer), two participants receiving 2 × Valneva (3200 IU/day offer), eight participants receiving a combination of vaccines (3200 IU/offer: n=3; 800 IU/day offer: n=3; no offer: n=2), and one participant unknown vaccine types (no offer). (3) Missing values: 25(OH)D concentration missing for three participants in 3200 IU/day arm and two participants in 800 IU/day arm. (4) Baseline 25(OH)D not determined for participants randomly assigned to the no offer arm.

### **Table S2.** Baseline characteristics of participants contributing data to analysis of neutralising antibody titres and cellular responses (sub-study 3), by allocation

|  | **Overall (n=100)** | **No offer (n=31)** | **800 IU/day offer (n=29)** | **3200 IU/day offer (n=40)** |
| --- | --- | --- | --- | --- |
| Age, years |  |  |  |  |
| Median (IQR) | 66.5 (61.4–68.9) | 66.5 (59.7–69.5) | 66.6 (64.2–69.0) | 66.4 (62.0–68.3) |
| Range | 16.6–87.2 | 18.4–82.8 | 26.8–83.1 | 16.6–87.2 |
| Sex |  |  |  |  |
| Female | 59 (58.4%) | 15 (48.4%) | 19 (65.5%) | 25 (62.5%) |
| Male | 41 (41.0%) | 16 (51.6%) | 10 (34.5%) | 15 (37.5%) |
| Ethnicity |  |  |  |  |
| White | 96 (96.0%) | 30 (96.8%) | 27 (93.1%) | 39 (97.5%) |
| Asian/Asian British | 1 (1.0%) | 0 | 0 | 1 (2.5%) |
| Black/African/Caribbean/Black British | 0 | 0 | 0 | 0 |
| Mixed/Multiple/Other | 3 (3.0%) | 1 (3.2%) | 2 (6.9%) | 0 |
| Body-mass index, kg/m² |  |  |  |  |
| <25 | 45 (45.0%) | 15 (48.4%) | 13 (44.8%) | 17 (42.5%) |
| 25–30 | 44 (44.0%) | 15 (48.4%) | 11 (37.9%) | 18 (45.0%) |
| >30 | 11 (11.0%) | 1 (3.2%) | 5 (17.2%) | 5 (12.5%) |
| Self-assessed general health |  |  |  |  |
| Excellent | 26 (26.0%) | 7 (22.6%) | 7 (24%) | 12 (30.8%) |
| Very good | 39 (39.0%) | 10 (32.3%) | 13 (45%) | 16 (41.0%) |
| Good | 28 (28.0%) | 10 (32.3%) | 8 (28.0%) | 10 (25.6%) |
| Fair | 5 (5.0%) | 4 (12.9%) | 0 | 1 (2.6%) |
| Poor | 1 (1.0%) | 0 | 1 (3.0%) | 0 |
| Medically diagnosed disease |  |  |  |  |
| Hypertension | 27 (27.0%) | 7 (22.6%) | 9 (31.0%) | 11 (27.5%) |
| Diabetes | 8 (8.0%) | 3 (9.7%) | 0 | 5 (12.5%) |
| Heart disease | 3 (3.0%) | 1 (3.2%) | 0 | 2 (5.0%) |
| Asthma | 14 (14.0%) | 4 (12.9%) | 7 (24.1%) | 3 (7.5%) |
| COPD | 2 (2.0%) | 1 (3.2%) | 1 (3.4%) | 0 |
| Previous SARS-CoV-2 infection^(1)^ | 2 (2.0%) | 0 | 0 | 2 (5.0%) |
| Type of vaccine administered, primary course |  |  |  |  |
| 2 × ChAdOx1 | 67 (67.0%) | 22 (71.0%) | 22 (75.9%) | 23 (57.5%) |
| 2 × BNT162b2 | 33 (33.0%) | 9 (29.0%) | 7 (24.1%) | 17 (42.5%) |
| Other | 0 | 0 | 0 | 0 |
| Inter-dose interval, days | 76 (70–78) | 75 (70–78) | 77 (70–80) | 76 (70–78) |
| Interval from second vaccine dose to blood sample, days | 82 (69–103) | 82 (65–93) | 81 (65–101) | 83 (71–125) |
| Mean 25(OH)D, nmol/L (SD) [range]^(2)^ | 39.3 (13.1) | --^(2)^ | 38.2 (14.1) | 40.1 (12.4) |
| <25.0 | 8 (8.0%) | --^(2)^ | 4 (13.8%) | 4 (10.0%) |
| 25.0 to <50.0 | 48 (48.0%) | --^(2)^ | 21 (72.4%) | 27 (67.5%) |
| 50.0 to <75.0 | 13 (13.0%) | --^(2)^ | 4 (13.8%) | 9 (22.5%) |
| ≥75.0 | 0 | --^(2)^ | 0 | 0 |
| Not determined | 31 (31.0%) | 31 (100.0%) | 0 | 0 |

Data are n (%) or median (IQR) unless specified otherwise. Abbreviations: IQR, interquartile range; SD, standard deviation; 25(OH)D, 25-hydroxyvitamin D. (1) Reported test-positive infection before provision of blood sample. (2) Baseline 25(OH)D not determined for participants randomly assigned to the no-offer arm.

### **Table S3:** Dichotomous outcomes by allocation

|  | **No offer** | **800 IU/day offer** | **3200 IU/day offer** | **Either offer** | **800 IU/day *vs* no offer** | | **3200 IU/day *vs* no offer** | | **Either offer *vs* no offer** | |
| --- | --- | --- | --- | --- | --- | --- | --- | --- | --- | --- |
|  |  |  |  |  | Adjusted OR (95% CI) | P | Adjusted OR (95% CI) | P | Adjusted OR (95% CI) | P |
| Proportion with breakthrough SARS-CoV-2 infection,  sub-study 1 | 49/908 (5.4%) | 66/944 (7.0%) | 59/956 (6.2%) | 125/1900 (6.6%) | 1.34 (0.91 to 1.97)* | 0.141 | 1.19 (0.80 to 1.76)* | 0.392 | 1.26 (0.89 to 1.77)* | 0.192 |
| Proportion of participants with detectable anti-S IgGAM post vaccination,  sub-study 2 | 533/555 (96.0%) | 628/646 (97.2%) | 638/652 (97.9%) | 1266/1298 (97.5%) | 1.29 (0.64 to 2.60)** | 0.474 | 1.66 (0.80 to 3.42)** | 0.172 | 1.42 (0.78 to 2.59)** | 0.248 |

Abbreviations: IQR, interquartile range; OR, odds ratio. *Adjusted for age, sex, educational attainment, frontline worker status, number of people per bedroom, schoolchildren (5–15 years) at home with participant, primary vaccination course, previous SARS-CoV-2 infection, season of first vaccination, inter-dose interval, and use of anticholinergics. **Adjusted for age, sex, body-mass index, days from second vaccine dose to DBS sample, general health, season of second vaccination, inter-dose interval, primary vaccination course, and previous SARS-CoV-2 infection.

### **Table S4:** Continuous immunological outcomes by allocation: intervention arms pooled

|  | **No offer** | **Either offer** | **Adjusted % difference for either offer vs. no offer (95% CI)*** | **P†** |
| --- | --- | --- | --- | --- |
| **Sub-study 2** | | | | |
| Anti-S IgGAM ratio | 2.8 (1.9 to 3.9) [n=555] | 2.9 (1.9 to 4.1) [n=1298] | -46.6% (-85.6 to 98.0) | 0.344 |
| **Sub-study 3** | | | | |
| Neutralising antibody titre | 186.9 (119.8 to 406.7) [n=29] | 237.6 (81.2 to 492.3) [n=66] | -2.4% (-39.5 to 57.5) | 0.920 |
| S peptide-stimulated IFN-γ in whole blood supernatant, ng/mL | 0.011 (0.003 to 0.025) [n=29] | 0.013 (0.005 to 0.037) [n=65] | 59.1% (-22.2 to 225.3) | 0.200 |
| S peptide-stimulated TNF in whole blood supernatant, ng/mL | 0.000 (0.000 to 0.003) [n=29] | 0.000 (0.000 to 0.005) [n=67] | -45.1% (-78.1 to 37.5) | 0.197 |
| S peptide-stimulated IL-6 in whole blood supernatant, ng/mL | 0.136 (0.012 to 0.719) [n=28] | 0.125 (0.000 to 1.909) [n=66] | -48.2% (-88.1 to 125.0) | 0.375 |
| S peptide-stimulated CXCL-8 in whole blood supernatant, ng/mL | 0.790 (0.254 to 2.563) [n=26] | 0.909 (0.371 to 5.917) [n=66] | 13.7% (-61.1 to 233.0) | 0.812 |
| LPS-stimulated IFN-γ in whole blood supernatant, ng/mL | 0.020 (0.004 to 0.067) [n=29] | 0.028 (0.007 to 0.072) [n=65] | 29.7% (-46.5 to 214.3) | 0.561 |
| LPS-stimulated TNF in whole blood supernatant, ng/mL | 0.542 (0.465 to 0.919) [n=29] | 0.513 (0.319 to 0.923) [n=68] | -37.2% (-72.7 to 44.4) | 0.270 |
| LPS-stimulated IL-6 in whole blood supernatant, ng/mL | 54.768 (32.362 to 73.147) [n=29] | 44.478 (30.651 to 81.109) [n=68] | 3.1% (-27.6 to 46.8) | 0.865 |
| LPS-stimulated CXCL-8 in whole blood supernatant, ng/mL | 4.231 (2.322 to 6.373) [n=28] | 2.642 (2.072 to 4.341) [n=68] | -20.2% (-44.2 to 14.2) | 0.214 |
| Percentage of S peptide-stimulated CD3+CD4+ cells positive for intracellular IFN-γ | 0.00% (0.00 to 0.02) [n=29] | 0.00% (0.00 to 0.02) [n=64] | -23.1% (-53.4 to 26.9) | 0.299 |
| Percentage of CD3-stimulated CD3+CD4+ cells positive for intracellular IFN-γ | 0.04% (0.00 to 0.12) [n=29] | 0.10% (0.02 to 0.39) [n=63] | 193.8% (11.0 to 677.8) | 0.030 |
| Percentage of S peptide-stimulated CD3+CD8+ cells positive for intracellular IFN-γ | 0.00% (0.00 to 0.04) [n=29] | 0.00% (0.00 to 0.06) [n=64] | 18.9% (-40.6 to 138.0) | 0.621 |
| Percentage of CD3-stimulated CD3+CD8+ cells positive for intracellular IFN-γ | 0.62% (0.18 to 2.15) [n=29] | 0.95% (0.26 to 2.09) [n=63] | 76.3% (-22.4 to 300.6) | 0.173 |
| Percentage of S peptide-stimulated CD3+CD4+ cells positive for intracellular IL-2 | 0.00% (0.00 to 0.02) [n=29] | 0.01% (0.00 to 0.02) [n=64] | 26.8% (-11.7 to 81.9) | 0.195 |
| Percentage of CD3-stimulated CD3+CD4+ cells positive for intracellular IL-2 | 0.12% (0.09 to 0.25) [n=29] | 0.18% (0.07 to 0.35) [n=63] | 51.3% (-13.7 to 165.5) | 0.146 |
| Percentage of S peptide-stimulated CD3+CD8+ cells positive for intracellular IL-2 | 0.00% (0.00 to 0.00) [n=29] | 0.00% (0.00 to 0.00) [n=64] | 21.4% (-17.7 to 78.9) | 0.323 |
| Percentage of CD3-stimulated CD3+CD8+ cells positive for intracellular IL-2 | 0.13% (0.05 to 0.26) [n=29] | 0.18% (0.08 to 0.41) [n=63] | 60.6% (-13.1 to 196.8) | 0.129 |
| Percentage of S peptide-stimulated CD3+CD4+ cells positive for intracellular TNF | 0.00% (0.00 to 0.00) [n=29] | 0.00% (0.00 to 0.02) [n=64] | 36.1% (-24.2 to 144.3) | 0.297 |
| Percentage of CD3-stimulated CD3+CD4+ cells positive for intracellular TNF | 0.37% (0.19 to 0.67) [n=29] | 0.73% (0.24 to 1.31) [n=63] | 90.0% (-3.7 to 275.1) | 0.064 |
| Percentage of S peptide-stimulated CD3+CD8+ cells positive for intracellular TNF | 0.02% (0.00 to 0.06) [n=29] | 0.00% (0.00 to 0.10) [n=64] | -31.9% (-70.1 to 54.8) | 0.354 |
| Percentage of CD3-stimulated CD3+CD8+ cells positive for intracellular TNF | 1.45% (0.72 to 3.84) [n=29] | 2.48% (1.05 to 4.40) [n=63] | 135.2% (4.9 to 427.3) | 0.038 |
| Percentage of CD3+CD4+ cells with naive phenotype | 35.8% (21.7 to 48.7) [n=29] | 36.5% (21.5 to 42.9) [n=64] | -3.6% (-22.4 to 19.7) | 0.737 |
| Percentage of CD3+CD8+ cells with naive phenotype | 32.5% (22.9 to 42.6) [n=29] | 24.6% (19.6 to 34.9) [n=64] | -13.7% (-28.9 to 4.7) | 0.134 |
| Percentage of CD3+CD4+ cells with central memory phenotype | 33.5% (26.6 to 40.6) [n=29] | 36.5% (30.8 to 46.1) [n=64] | 18.5% (1.1 to 39.1) | 0.037 |
| Percentage of CD3+CD8+ cells with central memory phenotype | 20.5% (13.9 to 28.8) [n=29] | 23.0% (14.1 to 30.5) [n=64] | 19.7% (-8.8 to 57.2) | 0.192 |
| Percentage of CD3+CD4+ cells with effector memory phenotype | 11.7% (7.3 to 15.7) [n=29] | 13.6% (9.5 to 17.8) [n=64] | 13.2% (-9.5 to 41.4) | 0.274 |
| Percentage of CD3+CD8+ cells with effector memory phenotype | 8.2% (5.6 to 12.7) [n=29] | 12.0% (7.1 to 15.1) [n=64] | 34.3% (2.8 to 75.4) | 0.031 |
| Percentage of CD3+CD4+ cells with EMRA phenotype | 3.9% (2.4 to 7.9) [n=29] | 3.4% (2.2 to 5.8) [n=64] | -32.4% (-53.6 to -1.6) | 0.041 |
| Percentage of CD3+CD8+ cells with EMRA phenotype | 14.9% (10.3 to 29.7) [n=29] | 19.6% (11.2 to 27.6) [n=64] | 7.8% (-21.3 to 47.7) | 0.638 |

Data are median (IQR) [n] unless otherwise specified. Values below the limit of detection are presented as 0. *Adjusted for age, sex, ethnicity, body-mass index, days from second vaccine dose to DBS sample, general health, inter-dose interval, and primary vaccination course. †Correction for multiple testing using the Benjamin and Hochberg procedure provides a critical p value of 0.0017. EMRA, terminally differentiated effector memory cells re-expressing CD45RA.

### **Table S5:** Sensitivity analysis: vaccine efficacy analysis in participants who did not report previous SARS-CoV-2 infection

|  | **No offer** | **800 IU/day offer** | **3200 IU/day offer** | **Either offer** | **800 IU/day *vs* no offer** | | **3200 IU/day *vs* no offer** | | **Either offer *vs* no offer** | |
| --- | --- | --- | --- | --- | --- | --- | --- | --- | --- | --- |
|  |  |  |  |  | Adjusted HR (95% CI) | P | Adjusted HR (95% CI) | P | Adjusted HR (95% CI) | P |
| Median days to breakthrough SARS-CoV-2 infection (IQR) | 123 (100 to 145) | 127 (90 to 146) | 103 (58 to 139) | 117 (73 to 145) | 1.27 (0.88 to 1.83) | 0.208 | 1.16 (0.80 to 1.69) | 0.427 | 1.23 (0.89 to 1.70) | 0.221 |

### **Table S6:** Sensitivity analysis: dichotomous outcomes by allocation in participants who did not report previous SARS-CoV-2 infection

|  | **No offer** | **800 IU/day offer** | **3200 IU/day offer** | **Either offer** | **800 IU/day *vs* no offer** | | **3200 IU/day *vs* no offer** | | **Either offer *vs* no offer** | |
| --- | --- | --- | --- | --- | --- | --- | --- | --- | --- | --- |
|  |  |  |  |  | Adjusted OR (95% CI) | P | Adjusted OR (95% CI) | P | Adjusted OR (95% CI) | P |
| Proportion with breakthrough SARS-CoV-2 infection,  sub-study 1 | 48/870 (5.5%) | 65/889 (7.3%) | 59/928 (6.4%) | 124/1817 (6.8%) | 1.34 (0.91 to 1.99)* | 0.136 | 1.18 (0.80 to 1.76)* | 0.402 | 1.27 (0.90 to 1.79)* | 0.175 |
| Proportion of participants with detectable anti-S IgGAM post vaccination,  sub-study 2 | 514/536 (95.9%) | 593/611 (97.1%) | 617/631 (97.8%) | 1219/1242 (98.1%) | 1.32 (0.66 to 2.65)** | 0.439 | 1.75 (0.85 to 3.60)** | 0.129 | 1.49 (0.82 to 2.69)** | 0.189 |

Abbreviations: IQR, interquartile range; OR, odds ratio. *Adjusted for age, sex, educational attainment, frontline worker status, number of people per bedroom, schoolchildren (5–15 years) at home with participant, primary vaccination course, previous SARS-CoV-2 infection, season of first vaccination, inter-dose interval, and use of anticholinergics. **Adjusted for age, sex, body-mass index, days from second vaccine dose to DBS sample, general health, season of second vaccination, inter-dose interval, primary vaccination course, and previous SARS-CoV-2 infection.

### **Table S7:** Sensitivity analysis: continuous immunological outcomes in participants who did not report previous SARS-CoV-2 infection

|  | **No offer** | **800 IU/day offer** | **3200 IU/day offer** | **Either offer** | **800 IU/day offer vs. no offer** | | **3200 IU/day vs. no offer** | | **Either offer vs. no offer** | |
| --- | --- | --- | --- | --- | --- | --- | --- | --- | --- | --- |
|  |  |  |  |  | Adjusted % difference  (95% CI)* | P | Adjusted % difference  (95% CI)* | P | Adjusted % difference  (95% CI)* | P |
| Anti-S IgGAM ratio (sub-study 2) | 2.7 (1.9 to 3.8) [n=536] | 2.8 (1.9 to 3.9) [n=611] | 2.8 (1.9 to 4.0) [n=631] | 2.8 (1.9 to 4.0) [n=1242] | -64.4% (-92.9 to 78.5) | 0.204 | -59.9% (-91.5 to 88.9) | 0.242 | -66.3% (-89.8 to 11.4) | 0.074 |
| S peptide-stimulated IFN-γ in whole blood supernatant, ng/mL (sub-study 3) | 0.011 (0.003 to 0.025) [n=29] | 0.013 (0.005 to 0.046) [n=28] | 0.013 (0.005 to 0.024) [n=37] | 0.013 (0.005 to 0.037) [n=65] | 2.3% (-6.6 to 12.1) | 0.616 | -0.5% (-4.8 to 4.0) | 0.823 | 1.7% (-4.0 to 7.6) | 0.568 |

Data are median (IQR) [n] unless otherwise specified. Values below the limit of detection are presented as 0. *Adjusted for age, sex, ethnicity, body-mass index, pre-vaccination anti-S IgGAM, days from second vaccine dose to DBS sample, general health, inter-dose interval, and primary vaccination course.
